# Disentangling Pontine Fiber Geometry and Microstructure in ARSACS Using Advanced MRI

**DOI:** 10.64898/2026.05.20.26353196

**Authors:** Ilana R Leppert, Aziz Benbachir, Jennifer SW Campbell, Santiago Coelho, Sajjad Feizollah, Mark C Nelson, Bernard Brais, Sirio Cocozza, G Bruce Pike, Roberta La Piana, Christine L Tardif

## Abstract

1

**Background:** Autosomal recessive spastic ataxia of Charlevoix-Saguenay (ARSACS) is a genetic disease characterized by spasticity and ataxia which reflects involvement of the corticospinal tracts (CST) and cerebellum. The primary involvement of the middle cerebellar peduncles (MCP) and transverse pontine fibers (TPF) at the crossing with the CST, and their role in the pathophysiology of the disease, is currently debated.

**Objectives:** Advanced MRI techniques capable of isolating sub-voxel microstructural parameters can test the hypothesis that the MCP and TPF are abnormally large, compressing the CST at their crossing, and potentially impairing CST development.

**Methods:** Tract macro- and micro-structural properties, including axon and tract caliber, axon density and geometry, and myelin content were estimated from diffusion-relaxometry and magnetization transfer imaging. These features were analyzed along segments of the CST, MCP, and TPF of 9 patients and 9 age-matched controls.

**Results:** While the CST showed significant decreases in tract size, axon caliber, and myelination throughout its length compared to controls (p<0.01), the MCP and TPF were relatively unaffected. In our group, neither the MCP nor the pons were enlarged. The proximal MCP showed an increase in axon caliber.

**Conclusions:** The increase in fractional anisotropy and axon density towards the center of the TPF could be driven by geometric confounds related to differences in the relative sizes of the CST and TPF compared to controls. This highlights the importance of investigating tract-specific microstructural profiles, particularly in regions of geometric complexity. The findings confirm the involvement of the CST, with a relatively limited involvement of the MCP and TPF.

**Highlights:**

- Tract-based decrease in axon density in ARSACS patients estimated using diffusion-relaxometry
- Corticospinal tracts are significantly more affected than transverse pontine fibers
- Higher intra-axonal T2 (increase in axon caliber) in patient MCP not seen in TPF
- Tractometry helps resolve geometrical confounds that can cloud global tract measures

**Graphical Abstract:** 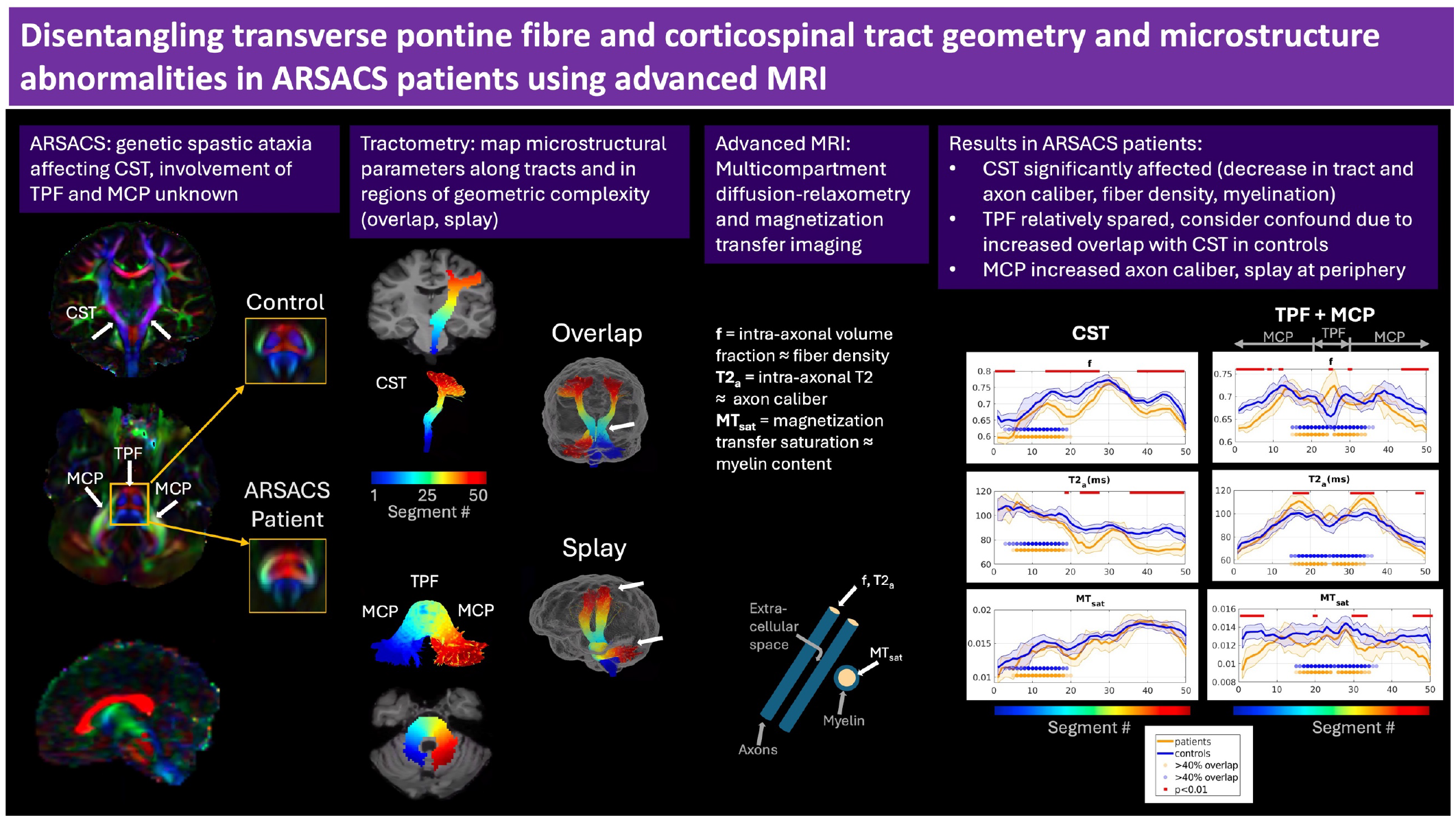

## 2 Introduction

Autosomal recessive spastic ataxia of Charlevoix-Saguenay (ARSACS, MIM 270550) is a hereditary disorder caused by biallelic pathogenic variants in the *SACS* gene. ARSACS is clinically characterized by early-onset spastic ataxia accompanied by peripheral neuropathy and retinal changes that appear to correspond to Retinal Nerve Fiber Layer thickening associated with aberrant accumulation of non-phosphorylated neurofilament H and glial fibrillary acidic protein. [1, 2]. Classic neuroradiological features include atrophy of the superior vermis, cerebellar hemispheres, and cervical spinal cord[3]. Supratentorial parietal cortical atrophy and thinning of the corpus callosum are also observed in more advanced stages of the disease. Linear pontine hypointense abnormalities are often observed in T2-weighted magnetic resonance images (MRI) and are thought to correspond to fibers of the corticospinal tract (CST) [4-6]. Several studies have qualitatively reported a thickening of the proximal middle cerebellar peduncles (MCP) as well as a ‘bulky’ pons [4, 5, 7, 8]. The latter finding was confirmed in a quantitative analysis of a large ARSACS cohort that reported a significant increase in the surface area of the pons in these patients [9].

Diffusion tensor imaging (DTI) [10], which is increasingly applied in clinical practice, uses a tensor to describe the diffusion of water along microstructurally oriented structures such as the axons in white matter. The fractional anisotropy (FA) describes the relative shape of this diffusion tensor and is often used as a measure of white matter integrity. However, it reflects a combination of intrinsic microstructural properties and voxel-scale fiber orientation organization. Previous DTI studies (reviewed in [11]) have observed an increase in FA in the pons of ARSACS patients relative to healthy controls [7], as well as an over-representation of diffusion tractography streamlines originating in the cerebella and entering the pons through the MCP and crossing the pons (transverse pontine fibers (TPF)). In contrast, the CST exhibited a decreased FA and a reduced number of reconstructed streamlines. This led to the hypothesis, raised by some studies, that abnormally large or excessively numerous TPF mechanically displace or compress the CST, potentially disrupting its normal developmental trajectory [4, 7].

As DTI relies on a single tensor, it cannot capture more complex geometries such as fibers crossing or kissing within a voxel. In these conditions, which occur in over 90% of white matter voxels in the human brain [12], the observed voxel average FA will be low despite the presence of highly directional and intact axons. This geometrical confound, can lead to misinterpretations, whereby changes in FA are indicative of changes in macroscopic white matter tract geometry rather than microstructure [13].

A recent study of one of the largest groups of ARSACS patients used advanced diffusion imaging and tractography techniques to address some of these geometrical confounds [14]. The results suggest that the MCP volume was increased while the TPF volume was decreased compared to controls. In addition, significant microstructural differences were reported in the CST (decrease in FA and neurite density index). Nonetheless, because of the complex tract geometry within the pons, including inferior-superior coursing CST and left-right TPF, it remains partially unanswered whether the observed differences in FA, neurite density, and tractography streamline counts reflect partial volume effects and macroscopic geometry, or true microstructural differences in the CST and TPF of ARSACS patients compared to controls.

To further disentangle macroscopic tract geometry from microstructural features, we performed an advanced multimodal quantitative MRI study of ARSACS patients. We investigated quantitative MRI parameters with varying sensitivity to intra- and extra-axonal water content, axon packing geometry, and myelin content, while being largely insensitive to macroscopic geometry.

## 3 Methods

### 3.1 Participants

Ten individuals with molecularly confirmed ARSACS were enrolled in the study. Nine (age = 36.3 ± 7.3 years; 5 males) completed the entire protocol. The age, sex, clinical SARA (Scale for the Assessment and Rating of Ataxia) scores, and radiological findings are detailed in Table 2. Seven of the 9 subjects (77.8%) carried the classical French-Canadian c.8844delT genetic variant. Nine age- and sex-matched healthy controls (age = 29 ± 7 years; 5 females) completed all scans. This study was conducted on data acquired at the Montreal Neurological Institute within the PROSPAX project (ClinicalTrials.gov no: NCT04297891), approved by the McGill University Health Center Research Ethic Committee. Written informed consent was obtained from each participant prior to enrollment.

### 3.2 Contrast Mechanisms

The MRI contrasts, their derivatives, and the interpretation of the various microstructural parameters included in this study are described below and summarized in Table 1.

**Table 1:** Parameters that can be estimated from the different acquisitions, and the microstructural features they are sensitive to.

| Estimated MRI Parameter | Compartment it's sensitive to | Voxel/orientation averaged | Orientation-specific | Microstructural feature index |
| --- | --- | --- | --- | --- |
| T2<sub>a</sub> | Intra-axonal relaxation time | x |  | Axon caliber |
| T2<sub>e</sub> | Extra-axonal relaxation time | x |  | Axon packing density |
| f | Intra-axonal volume | x |  | Fiber density |
| FA | Relative geometry of intra and extra-axonal spaces | x |  | Macroscopic anisotropy |
| MT<sub>sat</sub> | Membrane interfaces | x |  | Myelin content |
| AFD | Intra-axonal volume (T2-weighted) |  | x | Fiber density |

**Table 2:**
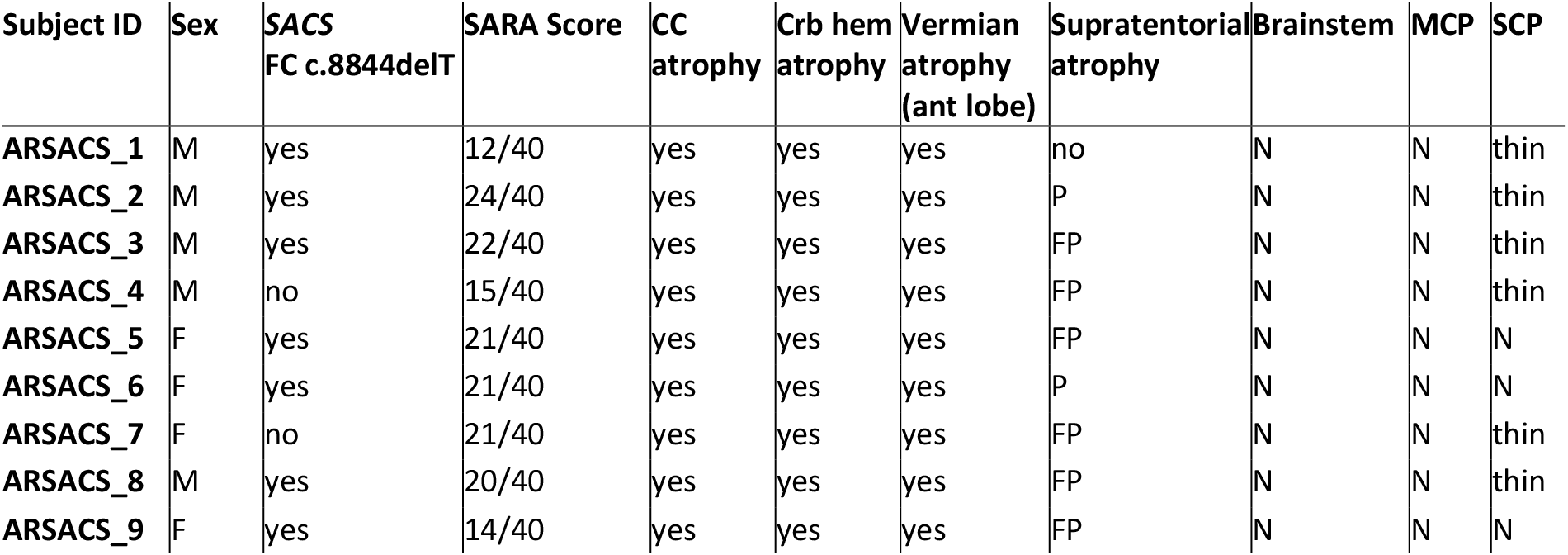
Demographic, clinical and radiological findings in the cohort of 9 subjects with ARSACS included in the study. Legend: SARA, Scale for the Assessment and Rating of Ataxia; CC, corpus callosum; Crb, cerebellar; Hem, hemispheric; MCP, middle cerebellar peduncle; SCP, superior cerebellar peduncle; F, frontal; P, parietal; N, normal.

| Subject ID | Sex | SACS<br>FC c.8844delT | SARA Score | CC<br>atrophy | Crb hem<br>atrophy | Vermian<br>atrophy<br>(ant lobe) | Supratentorial<br>atrophy | Brainstem | MCP | SCP |
| --- | --- | --- | --- | --- | --- | --- | --- | --- | --- | --- |
| ARSACS_1 | M | yes | 12/40 | yes | yes | yes | no | N | N | thin |
| ARSACS_2 | M | yes | 24/40 | yes | yes | yes | P | N | N | thin |
| ARSACS_3 | M | yes | 22/40 | yes | yes | yes | FP | N | N | thin |
| ARSACS_4 | M | no | 15/40 | yes | yes | yes | FP | N | N | thin |
| ARSACS_5 | F | yes | 21/40 | yes | yes | yes | FP | N | N | N |
| ARSACS_6 | F | yes | 21/40 | yes | yes | yes | P | N | N | N |
| ARSACS_7 | F | no | 21/40 | yes | yes | yes | FP | N | N | thin |
| ARSACS_8 | M | yes | 20/40 | yes | yes | yes | FP | N | N | thin |
| ARSACS_9 | F | yes | 14/40 | yes | yes | yes | FP | N | N | N |

### 3.2.1 Diffusion Tensor

Diffusion MRI measures the directional dependence of water diffusion, which arises from the organized microstructure of tissues. In white matter, the coherent arrangement of axons gives rise to anisotropic diffusion, with greater diffusivity along the dominant fiber orientation (D_∥_) than perpendicular to it (D_⊥_). DTI involves fitting a second-order tensor to the diffusion encoded MRI data, enabling the characterization of both the intensity of diffusion and its principal direction within a voxel [10]. FA combines D_∥_ and D_⊥_ to provide a measure of anisotropy, with 0 representing isotropic diffusion (D_∥_= D_⊥_) and 1 referring to diffusion along a single axis (D_⊥_ = 0). This metric is often used as a measure of white matter integrity, even though it reflects both microstructural properties and fiber orientation organization within the voxel.

### 3.2.2 Fiber orientation distribution

Applying diffusion encoding at higher diffusion weightings and angular resolution (typically 30-90 directions rather than the minimum of six required for DTI) helps better describe the diffusion displacement orientation distribution. Through deconvolution with a single fiber response function, this information can be used to estimate a fiber orientation distribution (FOD) within each voxel [15]. This allows the reconstruction of multiple fiber elements (called fixels) at different orientations within a voxel along with their relative contribution and organization (complexity [16]). The apparent fiber density (AFD), which is the integral of the FOD lobes, is thus orientation-specific and provides the relative intra-axonal volume that can be associated with each fibre population in a voxel [17].

### 3.2.3 Tractography

In diffusion tractography, streamlines are reconstructed by following the principal diffusion direction from voxel to voxel, which can then be combined into bundles representing plausible white matter fiber tracts [18]. Whereas DTI models a single fiber direction per voxel, fixel-based approaches that incorporate multiple orientations and anatomical priors [19] enable more reliable streamline reconstruction, particularly in regions of crossings where large tracts might obscure smaller ones, such as the TPF and CST.

### 3.2.4 Combined diffusion and relaxometry

Concurrently encoding multiple MRI contrast mechanisms in an image can provide further insight into microstructural features. By varying the combination of different acquisition parameters, the relative contributions and features of microstructural compartments can be probed and estimated. In the case of diffusion-relaxometry, the standard model for diffusion in white matter (SM) [20] can be used to dissociate intra and extra-axonal compartments [21, 22]. The intra-axonal compartment, modeled as a thin cylinder, is described by a relative volume fraction f, parallel diffusivity D_a_ and an intra-axonal relaxation time T2_a_. The extra-axonal compartment is described by a parallel D_e∥_ and perpendicular D_e⊥_ diffusivity, a volume fraction, f_e_, and a relaxation time T2_e_. Finally, the free water compartment, f_w_, is described by an isotropic diffusivity D_w_ and relaxation time T2_w_. The intra-axonal T2 (T2_a_) is associated with axon caliber, whereby smaller intra-cellular space leads to more membrane surface interactions and thus shorter T2 [23-25]. Estimating parameters from this model provides additional insight into the microstructural changes in the tracts of interest.

### 3.2.5 Magnetization transfer saturation

Magnetization transfer (MT) imaging is sensitive to the voxel-wise macromolecular content. In white matter, the largest contribution typically arises from myelin sheaths surrounding axons. Although several studies have shown that the magnetization transfer ratio (MTR) correlates with myelin-based protein histology (summarized through meta-analysis [26, 27]), its sensitivity and specificity is confounded by other factors such as edema and inflammation [28]. By adding an additional measurement, the magnetization transfer saturation (MT_sat_) can be used to reduce the effect of high-T1 components (such as edema) on MTR [29]. Note that due to its short T2 relaxation time, signal from water within the myelin sheath itself is invisible to the relaxometry and diffusion regimes explained above. Thus, in the context of this study, the aim was to estimate a complementary measure to determine whether changes in myelination could be detected in the TPF.

### 3.3 Acquisition protocol

Participants were scanned on a 3T Siemens Prisma-Fit system, equipped with a 32-channel head coil. T1-weighted anatomical magnetization-prepared rapid gradient echo (MPRAGE) images were acquired with the following parameters: 1 mm isotropic, TE/TI/TR = 2.98/900/2300 ms, FA = 8°. The following sequences were used for the computation of MT_sat_: (i) MP2RAGE with compressed-sensing [30] (1mm isotropic, TE/TI_1_/TI_2_/TR = 2.9/940/2830/5000 ms, FA=4°, 4.6x undersampling, 4min) (ii) transmit field B1 map [31] (2.5×2.5×3 mm^3^, 40s) (iii) a custom MT-weighted sequence with the following parameters: 1mm isotropic, TE/TR=2.42/27 ms, MT pulse (offset frequency = 2kHz, FA=220°, 4.096ms, 6:16 min). Diffusion-weighted images were acquired using a custom sequence [32] with free gradient waveforms with a range and combination of b-values (0-6 ms/ µ m_2_), directions (7-60), b-tensor shape (linear, planar and spherical) and echo times (TE=62, 80, 92, 130 ms), TR=4.6 s, using an optimized protocol [33] at 2 mm isotropic for a scan time of 30 min.

### 3.4 Analysis

The anatomical MPRAGE images of subjects with ARSACS and healthy controls were blindly reviewed by an experienced rater (RLP, with more than 10 years of experience) for the qualitative screening of radiological features known to be reported in ARSACS [5]: cerebellar and vermian atrophy, corpus callosum thinning, fronto-parietal atrophy, superior cerebellar peduncle thinning. The radiological features of the ARSACS cohort are reported in Table 2. No pathological features were documented in control subjects. Figure 1 illustrates some representative images of the subjects with various degrees of cerebellar, vermian, corpus callosum and lobar atrophy.

**Figure 1:**
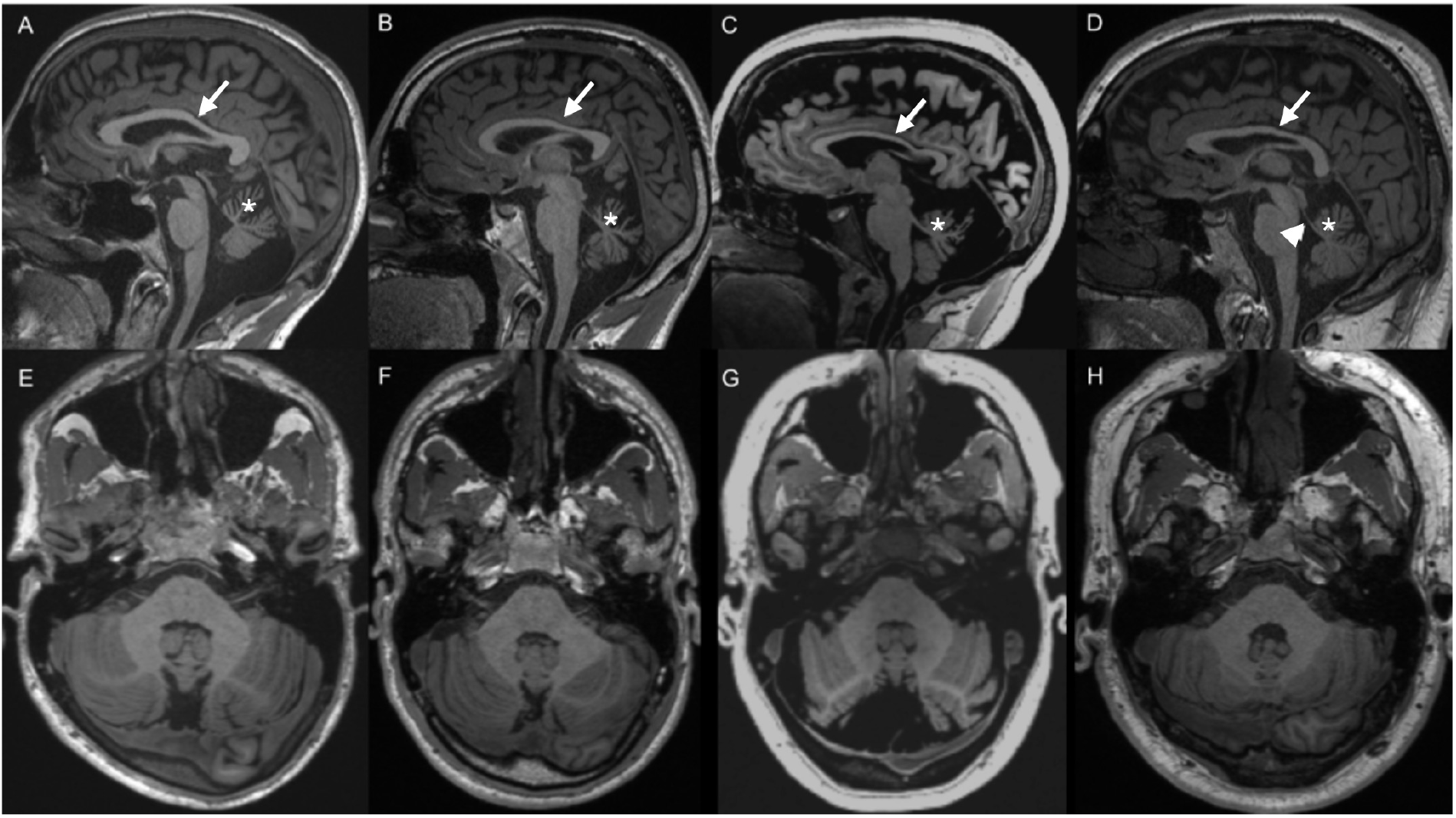
Representative MR images of subjects with ARSACS included in the study. Sagittal (A-D) and axial (E-H) MP2RAGE T1-weighted images showing various degrees of cerebellar, vermian (stars), corpus callosum (arrows) and lobar atrophy. A, E. Subject ARSACS_2 with vermian atrophy affecting particularly the anterior lobe and moderate corpus callosum thinning; B, F. Subject ARSACS_1 showing severe vermian atrophy, moderate corpus callosum and superior cerebellar peduncle thinning (arrowhead); C,G. Subject ARSACS_3 with severe vermian, corpus callosum and fronto-parietal atrophy; D, H. Subject ARSACS_8 with moderate vermian atrophy and thin corpus callosum.

The volumes of brain structures were derived from the anatomical MPRAGE data using FreeSurfer [34] and volBrain[35]). The diffusion-relaxometry data was preprocessed using the DESIGNER pipeline [36], which includes denoising [37], Gibbs ringing removal [38], and distortion correction and registration [39]. The preprocessed data were subsequently fit to the standard model for diffusion [22]. The high angular resolution linear b-tensor encoded portion of the diffusion-relaxometry dataset was used to compute FA, as well as to compute FODs using multi-shell multi-tissue deconvolution [40] followed by whole brain probabilistic tractography [41]. The AFD, complexity, and fixel count were all extracted from the FODs. Note that the AFD was computed per orientation and then corrected such that streamlines that are parallel in a voxel (i.e. same fixel) but belong to different tracts have distinct AFD values [42]. Finally, the MT_sat_ map was computed using the MT-weighted image and MP2RAGE T1map and included B1 correction [36].

The left (CST_L_) and right (CST_R_) CST and the combined MCP+TPF were delineated using anatomical priors from the Desikan-Killiany atlas [43] as endpoints and waypoints for the tractography streamlines. More specifically, the CST was defined by streamlines connecting the medulla to precentral cortex, while the MCP+TPF was defined by streamlines connecting the cerebellar cortices and passing through the pons. The MCP and TPF were considered separately because although diffusion tractography consistently generates streamlines that connect the cerebella through the pons, they do not correspond to an existing continuous anatomical pathway. In fact, the major cortico-ponto-cerebellar tracts course from the cortex to the contralateral cerebellar cortex, such that the TPF comprise the intersection of these bilateral pathways (diagram in Figure 2B). Because of the previously reported involvement of the cerebella, MCP, and TPF in ARSACS, analysis was limited to the inferior section of the pathways. The curvature of the streamlines was used to define the TPF as the portion of the tract running laterally, and the MCP as the portion of coherent streamlines connecting to the cerebellar cortices. This is further described in the following section.

**Figure 2:**
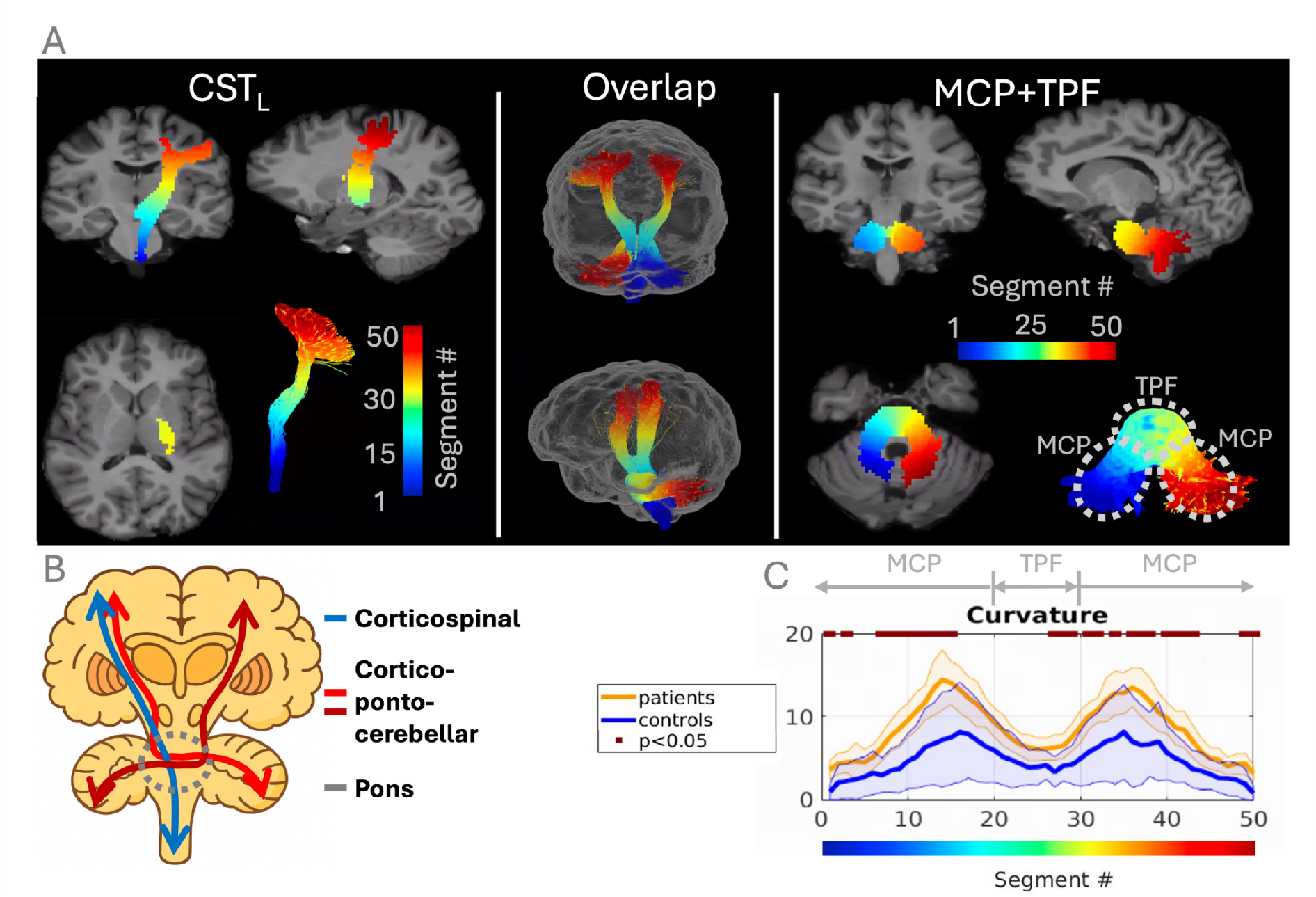
(A)Depiction of the delineation and division into 50 segments for the left cortical spinal tract (CST_L_) and the combined middle cerebellar peduncle (MCP) and transverse pontine fibers (TPF). The center panel depicts the overlap of the tracts. (B) Diagram of the bilateral cortico-ponto-cerebellar and corticospinal tracts. (C) The curvature of the MCP+TPF pathway. The change in curvature is used to delineate the sections: TPF corresponds to sections 20-30 and the MCPs correspond to the sections that connect the cerebella and the TPF (1-19 and 31-50).

Tractometry, or profile analysis, was used to map quantitative parameters onto streamlines and to summarize these values along segments of each tract [44]. This is especially interesting when investigating how MR-derived features are affected by crossing bundles, partial volume effects and changes in geometry and curvature. Using TractoFlow [45], the CST and MCP+TPF bundles were divided into 50 segments based on their centroid, as shown in Figure 2A. The curvature of the MCP+TPF is shown in Figure 2C. Although there are global differences in curvature between patients and controls (p<0.05), MCP and TPF were differentiated based on the change in curvature along the pathway, such that the TPF was defined as sections 20-30 and the MCP bilaterally (1-19 and 31-50).

Group differences in all measures were first assessed using an ANOVA that included age and sex as covariates (p<0.05). Subsequently, for the volumetric data, linear models were fit to test whether group differences varied as a function of age and sex (threshold p<0.05). Finally, for the profile analysis, parameters were averaged across individual segments and differences between patients and controls were evaluated using t-tests with false discovery rate correction (p<0.05).

## 4 Results

### 4.1 ARSACS patients show reduced brain size, but the MCP and pontine fibers are relatively spared

As shown in Figure 3, the intracranial volume (ICV) of ARSACS patients was smaller than that of controls (p=0.013, adjusted for age and sex). Furthermore, the cerebral white matter and cerebellum were significantly atrophic both in absolute terms and relative to the ICV (p=4.0e^-6^). Although the absolute volume of the pons was smaller in patients (p=0.05), it is not significantly different when normalized to ICV (p=0.14). The linear model revealed a significant group × age × sex interaction for the ICV (p=0.036). No other interactions were significant. In the tract-wise ICV-normalized analysis (boxplots in Figure 4A), the bilateral CST are significantly smaller (p=3.6e^-6^ and p=4.6e^-7^) while the volume occupied by the TPF is not increased in this cohort (p=0.14). The volume of each segment along the tracts (plot in Figure 4A based on segments in Figure 2A) sheds additional light on the source of these differences. In fact, although the CST tract volumes were significantly different throughout their length, the difference in the extent of lateral splay in the cortex (segments #35-45) was the most pronounced (Figure 4B).

**Figure 3:**
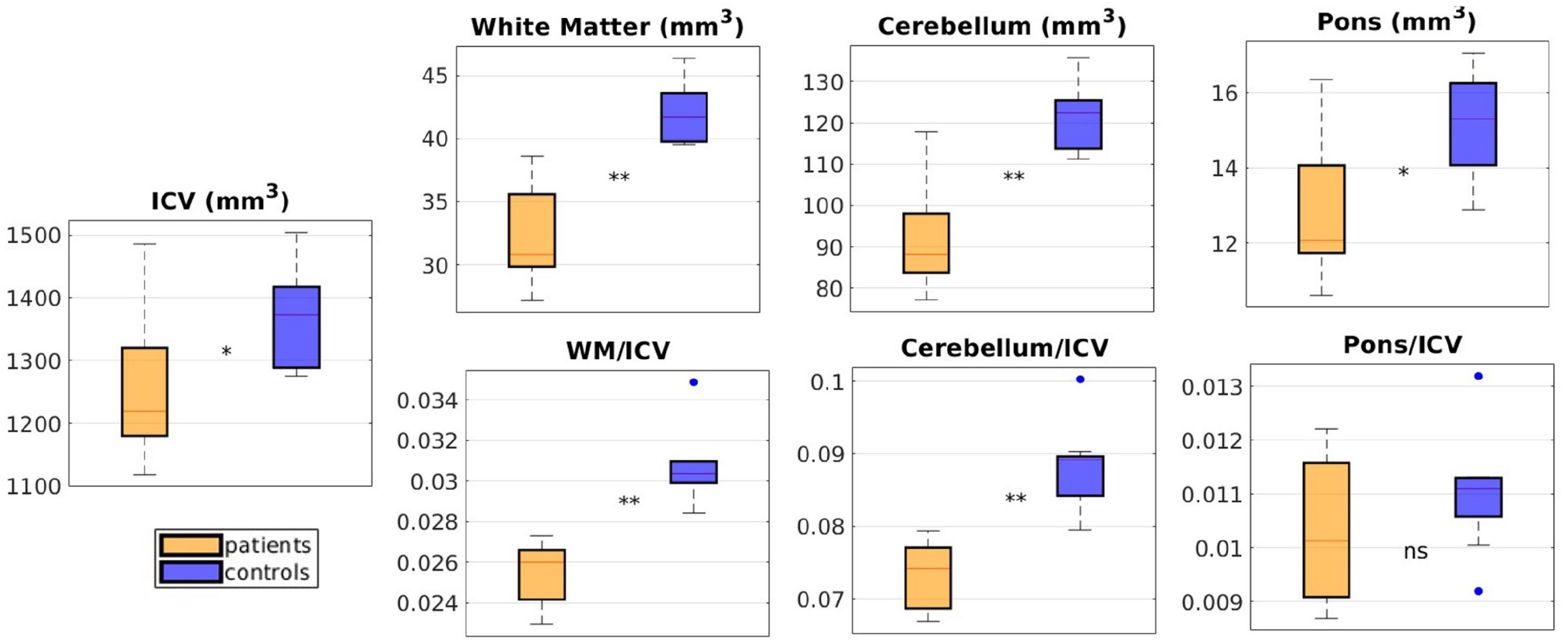
Anatomical volumes: absolute and normalized by intracranial volume (ICV). (**) significant at p<0.01, (*) significant at p<0.05, (ns) not significant.

**Figure 4:**
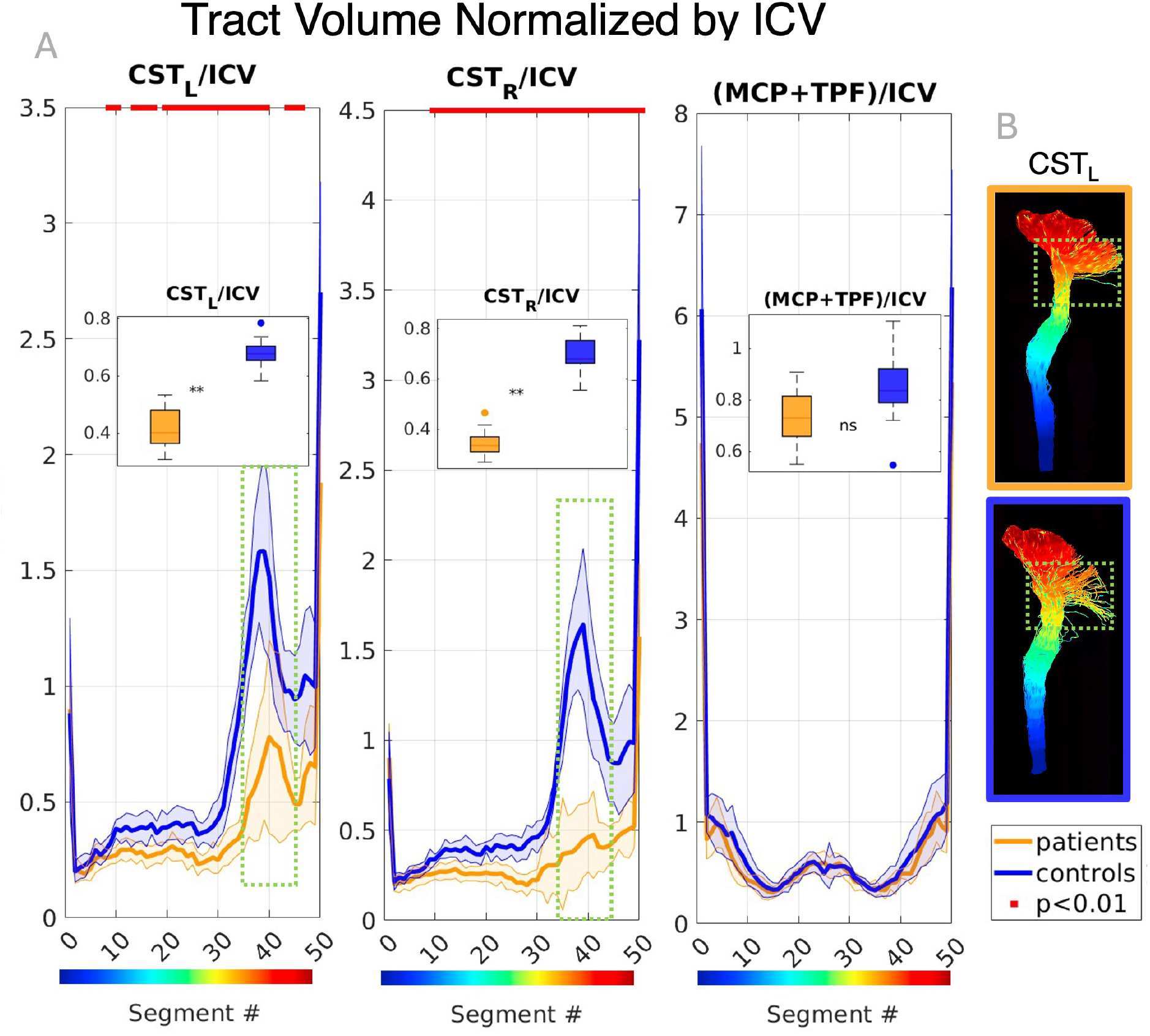
Tract volume normalized by intracranial volume (ICV): (A) Over the whole tract (boxplot) and along the 50 segments (line plot) of the left and right cortical spinal tracts (CST_L,_ CST_R_) and the combined middle cerebellar peduncles (MCP) and transverse pontine fibers (TPF). The red markers at the top of panel highlight segments that are significantly different (p<0.01) and the shaded area the standard deviation between subjects. On the boxplots: (**) significant at p<0.01 and (ns) not significant. (B) The dotted green boxes indicate where the CSTs differ the most: reduced lateral splay in the cortex of patients.

### 4.2 Widespread differences in CST microstructural parameters between ARSACS patients and controls

The bundle averages of each parameter are shown in Figure 5. All estimated parameters show a significant decrease in the CST of patients compared to controls (all p<0.01). For the TPF, there is a significant increase in FA (p=0.033) and decrease in MT_sat_ (p=0.040). In the MCP, there is a significant decrease in f (p=4.6e_-4_). There are no other significant differences within the TPF and MCP compared to controls.

**Figure 5:**
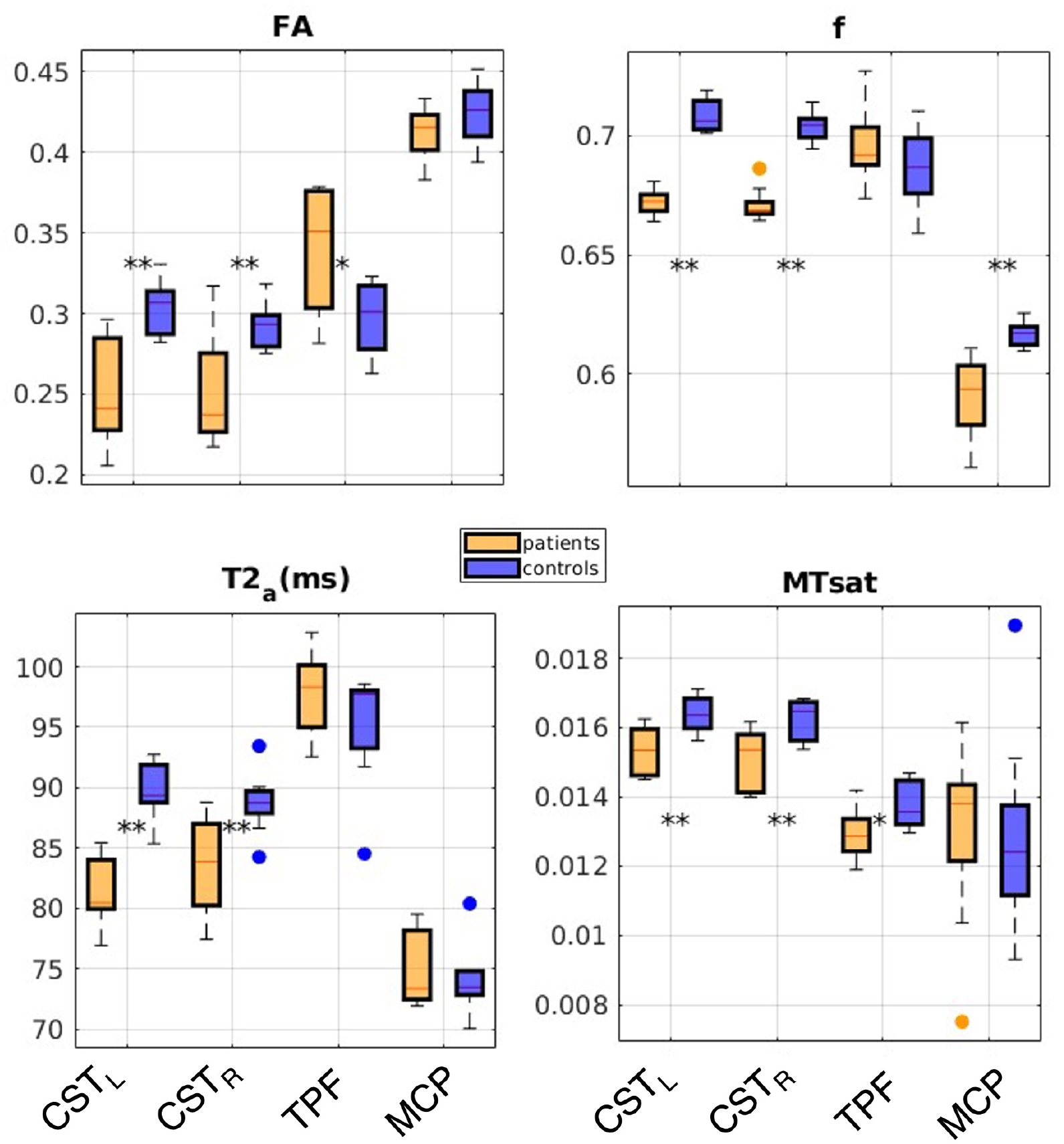
Whole-tract averaged parameters. (**) significant at p<0.01, (*) significant at p<0.05.

The profile analysis of these parameters in the CST_L_and MCP+TPF is shown in Figure 6 (CST_R_ shows a similar pattern and is included in supplementary Figure S1). The significantly different segments between patients and controls are highlighted in red. The blue and yellow dots below the profiles indicate where the CST and TPF overlap.

**Figure 6:**
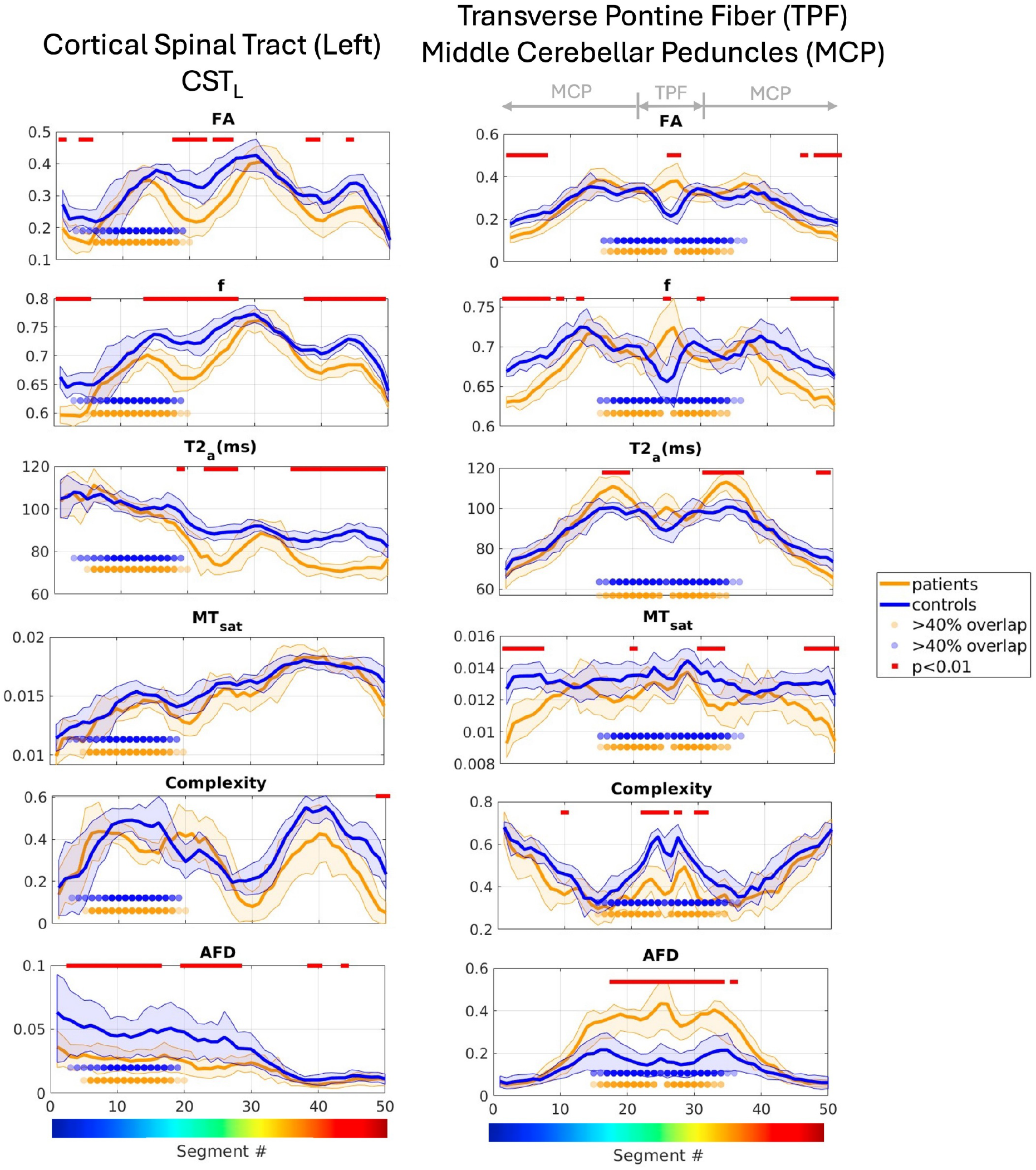
Microstructural profile analysis: Average patient and control profiles along segments of the cortical spinal tract (left) (CST_L_) and combined transverse pontine fibers (TPF) and middle cerebellar peduncles (MCP). The shaded regions correspond to the standard deviation between subjects, the shaded dots correspond to segments that overlap at greater than 40% between the two tracts, and red dots correspond to segments where patient and control profiles significantly differ (p<0.01). The right cortical spinal tract (CST_R_) shows similar results and is shown in the Supplementary Material.

For the CST profiles, the significant differences in FA, f and T2_a_ agree with the results in Figure 5. In contrast, despite being consistently lower in patients, MT_sat_ does not reach significance at the segment level. The f parameter shows more extensive differences than FA, especially in regions of high geometric complexity (i.e. segments 15-25 and 35-50). T2_a_ shows a similar pattern of significance inferior and superior to the internal capsule (segments 30-33). The inability to detect differences in the internal capsule is likely confounded by the presence of other tracts going through this region (resulting in a low AFD) and the fact that this range of segments contains both the medial and lateral portions of the tract. In segments where both the complexity is high and the CST and MCP+TPF overlap (i.e., the shaded dots corresponding to segments 5-20), there are no significant differences between tracts. Only the AFD, which is orientation-specific, can isolate the contributions aligned with the CST. This is significant for a large part of the tract, but no longer significant close to the cortex, where partial volume effects are exacerbated. Overall, the results in the CST of ARSACS patients point to a decrease in tract volume, intra-axonal volume (f), axon caliber (T2_a_), and myelin content (MT_sat_).

### 4.3 ARSACS patients have a higher intra-axonal T2 and decreased MT_sat_ in the MCP

The tract profiles show a significant increase in FA and f in the center of the TPF and decrease laterally towards the cerebellar cortices. The decrease in complexity due to relatively little overlap with the CST and elevated AFD in patients at the level of the TPF (center segments 20-30) supports reliability of the metrics in this region. In the periphery, near the cerebellar cortices, decreased AFD and increased complexity are indicative of partial volume effects. There is a noticeable increase in T2_a_ lateral to the TPF (i.e. in the MCP centered around segments 15 and 35) co-localized with a decrease in MT_sat_. Although there is consistent overlap with the CST in this area, in patients the AFD is high and the complexity low, indicating that the MCP are likely driving the result.

## 5 Discussion

Individuals with ARSACS exhibit significant atrophy of various cerebral and cerebellar structures compared to controls, particularly in the white matter. Although a recent study has shown an increase in the volume of the MCP[14], the volumetric analysis in our group points to the pons and the entire MCP+TPF being relatively spared. As confirmed by tract-based analysis, the size of the CST in ARSACS patients is significantly reduced throughout its length and exhibits reduced cortical splay compared to controls. The reduction in splay observed could also be attributed to lower detectability due to the tracts being small relative to the image resolution. The combination of unchanged MCP+TPF and smaller CST and cerebella could lead to the visual appearance of ‘bulky’ pons and MCP (e.g. [4, 8]). This difference in appearance is also likely driven by the significant increase in curvature in the MCP (Figure 2B).

Overall, in the tracts studied, there are significant differences across all microstructural parameters between ARSACS patients and controls. This is consistent with previous work that reported distributed decreases in FA [7, 46], and more specifically in the CST [14]. The f parameter from the Standard Model of diffusion in WM shows less variability than FA (as shown in Figure 5) because it reflects intra-axonal volume and is thus less affected by voxel-wise macroscopic geometry and partial volume effects [20]. Furthermore, MT_sat_ results indicate a general decrease in voxel-wise myelin content. The concurrent overall decrease in f, along with the increase in T2_e_ and D_e⊥_ (Figure S2) points to an increase in the size of the extra-axonal compartment and would be consistent with demyelination rather than hypomyelination. In other words, the diffusion measures are sensitive to packing density, which would be lower in the case of axonal loss or demyelination, compared to the case of tightly packed hypomyelinated fibers.

For the CST, the profiles highlight significant decreases along the tract length, such as reduced f and T2_a_, which reflect decreased intra-axonal content and potentially smaller axons, respectively [23-25]. This also aligns with previous CST tractometry work that showed similar FA profiles for patients and controls [14]. The lack of significance between patients and controls in some segments of the CST is likely due to extensive overlap with other tracts (either crossing or parallel) or partial volume effects near the cortex.

The profile analysis of the combined MCP+TPF reveals the importance of considering geometrical confounds when interpreting voxel-averaged microstructural parameters. Although there are significant decreases in FA, f and MT_sat_ towards the cerebellar cortex, the co-localized increase in complexity and decrease in AFD can confound the results and thus should not be overlooked. The relative increase in partial volume effects due to relatively smaller patient cerebella could also affect the results. Of particular interest is the significant increase in FA and f in the center of TPF of patients, where the streamlines are highly coherent (i.e. uni-directional), have a high volume fraction contribution (AFD), and a decrease in complexity. The increase in FA in the pons has been reported previously [4, 7, 8], but the underlying geometric variability between controls and patients make it difficult to interpret. In fact, the combination of the parameters reported here could be indicative of two different scenarios. On the one hand, it could be pointing to an increase in TPF fiber density in patients. Alternatively, a possible explanation is that parameters estimated in the TPF of controls are dominated by the CST, such that when its contribution is reduced in patients, the ‘true’, unaffected TPF parameters can be estimated. In other words, the changes reported in the pons could be driven by the changes in the CST rather than the TPF.

In the proximal portion of the MCP in patients, the observed increase in T2_a_ points to an increase in axon caliber. In this case, the complexity is similar for patients and controls, indicating that the results are likely not driven by relative changes in tract contribution.

Although first described in French-Canadian patients, ARSACS is now considered one of the most frequent recessive ataxias worldwide [47, 48]. For this reason, the relatively small sample size and homogeneity in terms of age, ethnicity, and SARA scores is a limitation of the current study. In future work, it will be important to verify whether our results can be replicated in larger cohorts with broader demographics. Also, longitudinal studies will clarify whether the observed findings include neurodevelopment components or reflect progressive neurodegeneration and demyelination.

In conclusion, our study extends the current understanding of the pathophysiology of ARSACS and furthers clarifies the significance of the CST and MCP+TPF involvements that have not been previously disentangled. These findings might be used to select imaging biomarkers to track disease evolution and the efficacy of future therapeutic strategies.

## Data Availability

All data produced in the present study are available upon reasonable request to the authors

## 6 Acknowledgments

We would like to thank Tobias Kober, Tom Hilbert and Gian Franco Piredda from Siemens Healthineers for use of the prototype compressed sensing MP2RAGE sequence. The custom MT-weighted and free waveform sequences were developed at the McConnell Brain Imaging Center and are available on the Siemens Teamplay website. Financial support was provided by the Ataxia Charlevoix-Saguenay Foundation (RLP, CLT), Brain Canada, Fonds de recherche du Québec-Santé (FRQS) Research Scholars (CLT, RLP), Killam Trusts (CLT), and Natural Sciences and Engineering Research Council (NSERC) Discovery Grant (CLT).

## 7 Author contributions

IRL (conceptualization, methodology, analysis, writing), AB (data curation, patient recruitment), JSWC (conceptualization, methodology, review and editing), SCoe (methodology, review), SF (methodology), MCN (data curation, review), BB (review), SCoc (methodology, review), GBP (review), RLP (conceptualization, funding, supervision, review), CLT (conceptualization, funding, supervision, review)

**Figure S1:**
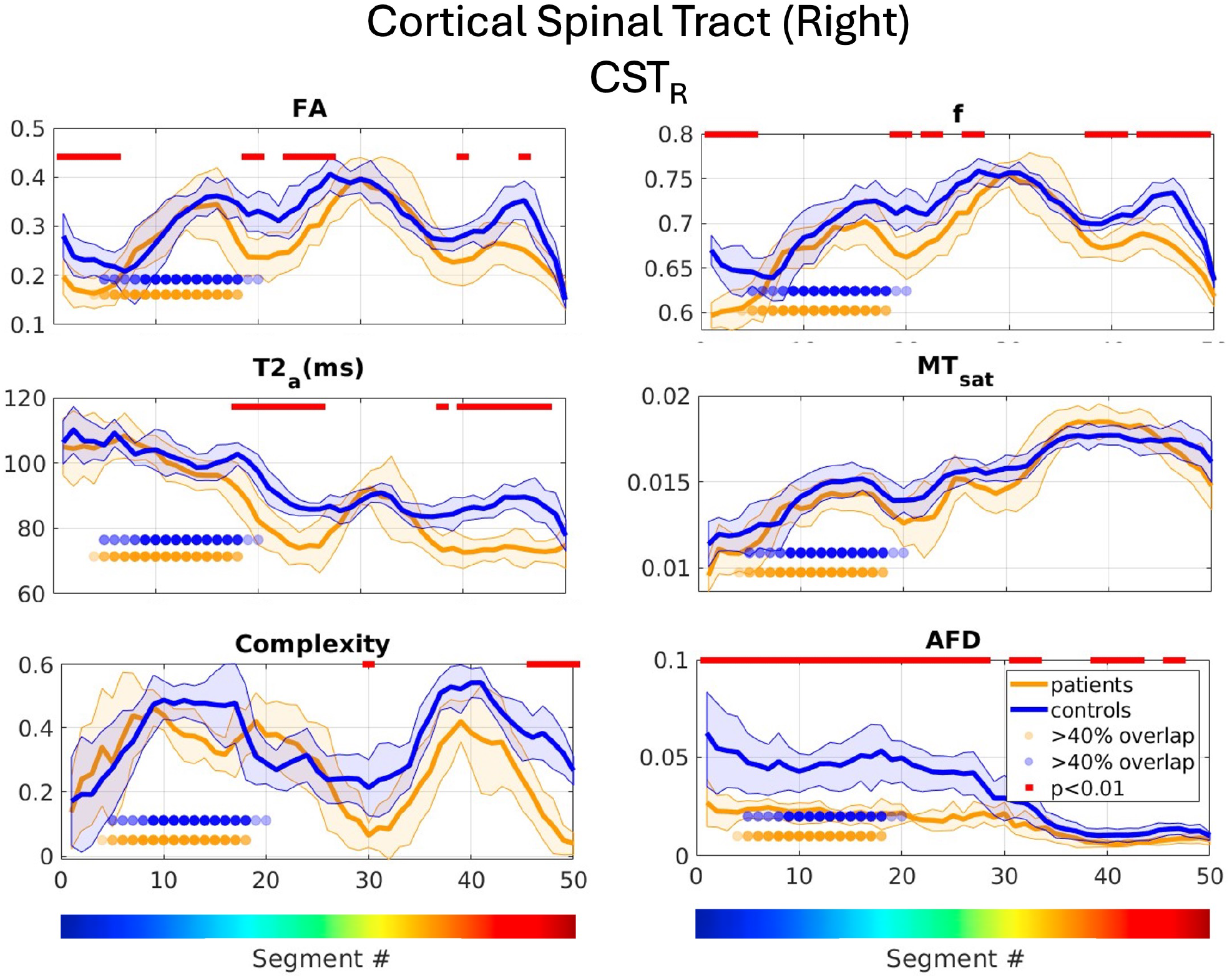
Microstructural profile analysis: Average patient and control profiles along segments of the cortical spinal tract (right) (CST_R_). The shaded regions correspond to the standard deviation between subjects, the shaded dots correspond to segments that overlap at greater than 40% between the two tracts, and red dots correspond to segments where patient and control profiles significantly differ (p<0.01).

**Figure S2:**
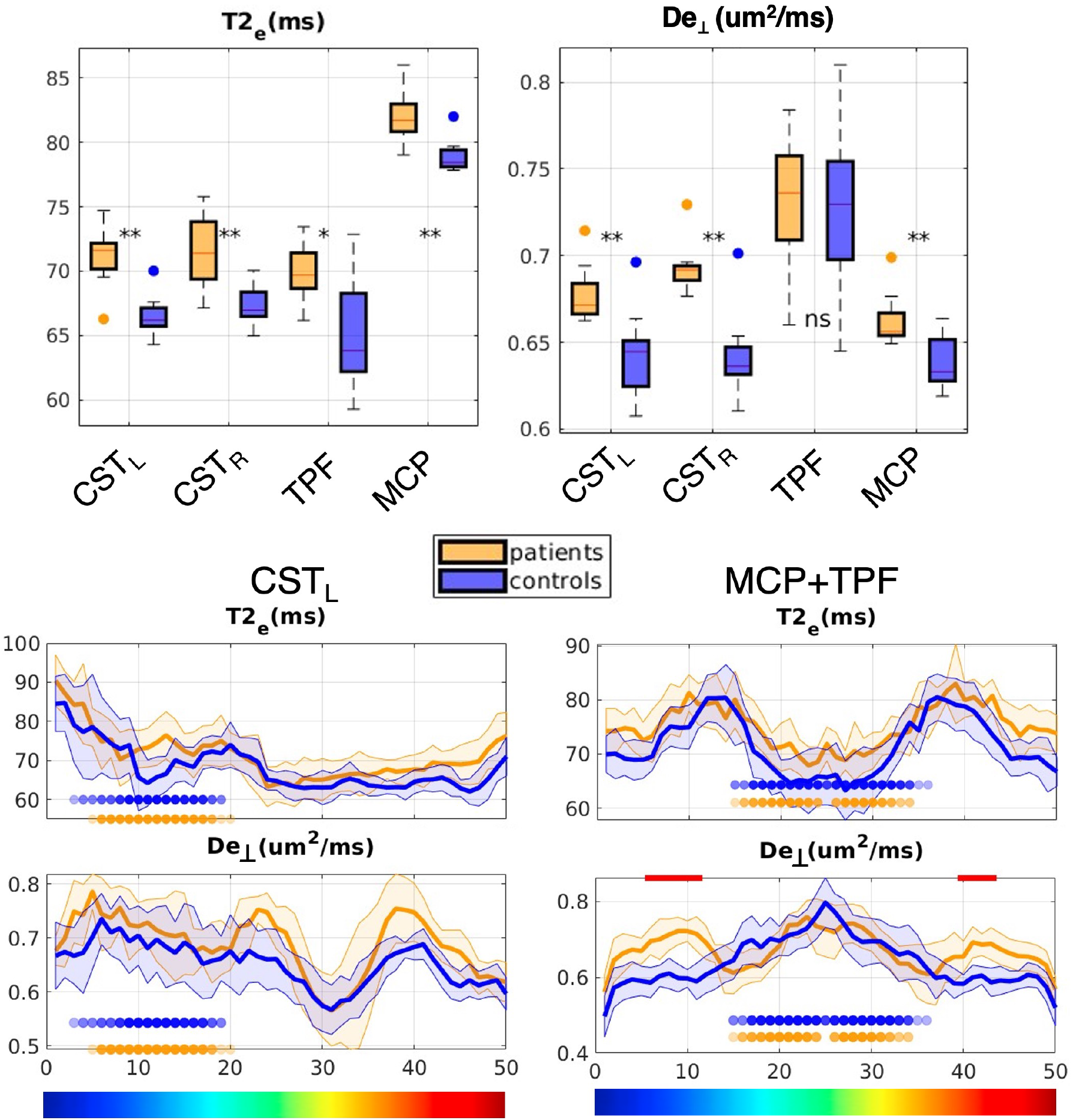
Whole-tract boxplots and profile analysis of T2_e_ and D_e⊥_ in the left (CST_L_) and right (CST_R_) cortical spinal tract, the middle cerebellar peduncles (MCP) and transverse pontine fibers (TPF). Boxplots: (**) significant at p<0.01, (*) significant at p<0.05, (ns) not significant and red dots correspond to segments where patient and control profiles significantly differ (p<0.01).

